# Applications of large language model in general practice: a qualitative interview study with professionals and patients

**DOI:** 10.64898/2026.10.30.26366911

**Authors:** Y. ter Heerdt, E.W.M.A. Bischoff, J. Yang, M. Timmers, J. Jung, R.C.A. van Linschoten

## Abstract

**Background:** Workload is increasing in general practice. Large language models (LLMs) are a promising tool that could relieve part of this workload. However, existing studies offer little insight into how professionals and patients want specific LLM applications to be implemented in practice.

**Objectives:** This study aimed to prioritize a broad range of LLM applications in general practice and identify the benefits, risks, and requirements of each application.

**Methods:** We conducted semi-structured one-on-one interviews with 12 general practitioners (GPs), 7 GP assistants, 6 practice nurses, and 6 patient representatives in the Netherlands between October and December 2025. To guide the interviews, we developed a list of 21 LLM applications derived from literature, existing tools, and GP input. Application were divided into 5 categories: administrative tasks, pre-consultation and triage, clinical decision support, management of (chronic) diseases, and patient interaction and communication. Participants discussed and prioritized each application. Transcripts were analyzed with a combined deductive and inductive approach.

**Results:** Priorities for LLM applications differed by group: GPs favored clinical decision support, assistants favored triage, and practice nurses and patient representatives favored chronic disease management. The most prioritized applications were summarizing consultations and triage and urgency classification (high priority 18/31; 58%), followed by clarification of medical information around the consultation (14/31; 45%). The qualitative analysis revealed three recurring considerations when applying LLMs for different tasks. First, participants questioned whether an LLM is needed, as simpler or existing tools may suffice. Second, they questioned whether LLMs relieve workload or create extra work. Third, a possible benefit on workload has to explicitly be weighed against risks such as mistakes, reduced personal care and accessibility, and negative effects on professionals.

**Conclusions:** Professionals and patients see potential for LLM support across many areas of general practice. Realizing this potential requires three steps: critically assessing whether an LLM is the right tool, establishing that it relieves workload, and designing applications so that their benefits outweigh the risks. Our application-level prioritization and analysis provide a starting point for assessment and design.

## Introduction

General practice is under increasing pressure [1]. In high-income countries, consultation rates and durations have risen steadily over recent years [2-4]. As a result, most general practitioners (GPs) experience a high workload; in the Netherlands, this was the case in 78% of practices in 2024 [5]. These high demands are associated with higher rates of emotional exhaustion [6], and negatively impact wellbeing of GPs [7]. Moreover, high workload and low wellbeing among physicians are associated with reduced quality and safety of care, and with poorer patient communication, adherence, and satisfaction [7-11].

Digital tools have the potential to play an important role in reducing pressure within general practices. Artificial intelligence (AI), especially large language models (LLMs), show promise in addressing common challenges such as administrative burden [12]. Recent studies have explored a wide range of LLM applications in general practice [13, 14]. Three main applications areas emerged: administrative support, clinical decision support, and patient support. Specific examples include automatically generating consultation notes [15], automatic clinical history taking [16], or translating medical information into accessible language for patients [17].

Several studies have investigated attitudes of GPs towards AI and LLMs. Most focus on AI broadly [18-21], while recent work addresses LLMs specifically [22, 23]. These studies address high-level themes on perceived benefits, risks, and implementation requirements. Possible benefits include reducing administrative work [21, 22] and supporting clinical decisions [19, 20, 23]. Concerns include accountability [18, 19, 22], threats to patient– practitioner relationships [20, 22], privacy [18, 23], accuracy and clinical reliability [18, 19, 21, 22], and deskilling [18, 19, 23]. Implementation requirements include integration into existing electronic health record (EHR) workflows [23], human oversight [22], and adequate clinician training [22]. A small number of studies have examined how future AI or LLM applications are prioritized in general practices [24, 25]. Participants in these studies saw value in both clinical (e.g., diagnostic support) and non-clinical applications (e.g., automated consultation notes) but generally favored non-clinical applications.

However, existing research offers limited guidance for implementing LLM tools in general practice for two reasons. First, most studies address broad, overarching themes rather than attitudes towards specific applications. As outlined above, LLMs applications in general practice span diverse applications [13, 14], each with its own trade-offs. Theme-level findings therefore offer little basis for deciding which tools to implement and how. Second, most studies include only GPs, overlooking other members of the practice team (e.g. GP assistants) as well as patients. Since these stakeholders are likely to be direct or indirect users of LLMs tools, their perspectives are needed for user-informed integration of LLMs into general practice [22].

To address these gaps, we conducted a semi-structured interview study with GPs, assistants, practice nurses, and patients in the Netherlands. Our aim was two-fold. First, we prioritized LLM applications across a broad range of general practice applications. Second, we analyzed the underlying stakeholder preferences and motivations at an application-specific level. These findings can inform future researchers and developers, enabling user-driven integration of LLMs within general practice.

## Methods

### Study design

This qualitative study examined LLM applications in general practice through semi-structured one-on-one interviews. We adopted a pragmatic, design-oriented approach aimed at generating insights to inform the development of LLM-based tools. This study was embedded in a broader public-private research project on LLM implementation in general practice, involving Erasmus Medical Centre (EMC), Technical University Delft (TU Delft) and Juvoly powered by Tandem Health (Dutch medical AI company).

### Study setting

The study was conducted in general practices across the Netherlands between October 2025 and December 2025. In the Netherlands, GPs act as gatekeepers and the first point of contact for patients. They solve the vast majority of health complaints, referring only 6.7% of contacts to secondary care, while accounting for around 5% of national healthcare expenditure [26]. General practice care is mainly delivered by three professionals: GPs *(huisarts)*, the main care providers; practice nurses (*praktijkondersteuners)*, who focus on chronic and mental health care; (GP) assistants *(doktersassistenten)*, the first point of contact for patients, responsible for triage and minor clinical procedures.

### Participants and recruitment

This study included two main participant groups. The first group consisted of general practice professionals: GPs, practice nurses and assistants. These would be the primary (professional) users of LLMs applications in Dutch general practices. The second group consisted of patient representatives, reflecting the perspective of patients as both users and recipients of LLM-supported care.

General practice professionals were recruited through the professional networks of the authors and national professional federations. We purposively oversampled GPs because, as the main care providers, they are the only stakeholders involved in all general practice areas. Patient representatives were recruited from regional general practice patient councils and from the Dutch Patient Federation. The latter had completed an AI training course provided by the federation. We selected representatives with either general practice or AI experience to ensure they could meaningfully engage with the hypothetical applications discussed.

### Interview materials and procedure

To guide the interview, we developed a list of 21 LLM applications in Dutch general practice. This predefined list allowed us to compare insights across interviews and elicit views on applications participants might not have raised spontaneously. We developed the application list from three sources: the literature on LLMs in general practice and other healthcare specialties, applications already available in practice, and suggestions from early conversations with GPs. We rephrased or merged applications to reduce overlap and ensure a similar level of abstraction. To enhance accessibility, we grouped the 21 applications into five categories spanning different areas of general practice. Finally, one GP reviewed the list for completeness and relevance. The full list is provided in Textbox 1.

The interviews consisted of two parts. In the first part, we discussed all 21 applications. In the second part, participants completed a prioritization exercise. They first selected any number of applications they considered high priority, followed by a top 3 selection from these high-priority applications. A more detailed interview protocol and guide is provided in Supplementary Files 1 and 2.

### Data collection

Prior to the interview, the following baseline characteristics were collected: age, gender, years of experience (general practice professionals only), self-reported LLM knowledge, and frequency of LLM use. Interviews were conducted face-to-face or via Microsoft Teams and lasted 45–60 minutes. During the interviews, applications were presented via printed cards (face-to-face) or a Miro board (online). All interviews were audio-recorded and automatically transcribed using Microsoft Teams. Transcripts were subsequently reviewed and manually corrected against the audio recording. During this process, all identifying information was removed from the transcript.

### Qualitative data analysis

The first author coded the interviews in MAXQDA (version 24.9.1), combining a deductive and an inductive approach. Deductively, each quote was assigned to the application it was related to. Within each application, quotes were structured into three subcodes: *benefits* (expected advantages of adoption), *risks* (potential negative consequences of adoption), and *requirements* (conditions for adoption). Where participants described multiple distinct functionalities within an application, we additionally double-coded the relevant quotes under a fourth subcode: *functionalities*, allowing sub-analysis per functionality. Inductively, quotes within each subcode were analyzed using thematic analysis across two coding cycles [27]. In the first cycle, we applied themeing the data [27] to summarize the content of each quote. In the second cycle, we applied pattern coding [27] to group similar codes and identify broader patterns

To ensure coding quality, the last author (RvL) was involved at multiple stages of the analysis as a second coder. First, the coding approach was developed in collaboration with RvL. Following the inductive coding cycles, RvL reviewed and validated the resulting code tree. To assess coding consistency, RvL independently coded one interview using the final code tree. Any discrepancies were discussed until consensus was reached.

### Researcher reflexivity

Interviews were conducted by the first author, a PhD candidate with a background in econometrics and no prior relationship with participants. The researcher’s training is more technically than clinically oriented, which may have shaped how interviews were conducted and analyzed. To broaden the interpretive perspective, both the interview design and the analysis were reviewed by the wider research team. This team incorporated design engineering, clinical (general practice), and industry backgrounds. Industry partners involved in the project had no role in data collection, analysis, or interpretation.

### Ethical considerations

The Human Research Ethics Committee of TU Delft approved this study on September 25, 2025 (application number 5969). Prior to inclusion, all participants provided written informed consent. All interview data were pseudonymized and stored on a secure server accessible only to the research team. Participants received remuneration of €100 (GPs) or €50 (other participants), reflecting differences in typical hourly income.

**Textbox 1**. Overview of the 21 potential LLM applications used in the interviews, grouped into five categories. Applications are numbered within each category according to their prioritization ranking (see Table 2).

**A.Administrative Tasks**

*A1. Summarizing consultations:* Summarizing doctor-patient conversations into structured formats such as SOAP.

*A2. Summarizing patient records:* Extracting relevant information from patient records to generate general or complaint-oriented summaries.

*A3. Processing medical letters -* Analyzing specialist letters and discharge reports to summarize their content and identify follow-up actions for the general practice.

*A4. Writing referral letters -* Drafting referral letters to other healthcare providers.

*A5. Organizing inbox and correspondence -* Sorting and prioritizing incoming messages (e.g., specialist letters, lab results) and flagging items that require immediate follow-up.

*A6. Coding of consultations -* Assigning medical codes, (e.g., ICPC, SNOMED-CT) and creating episodes of care based on consultations, specialist letters, or existing patient records.

**B. Pre-consultation and triage**

*B1. Triage and urgency classification* - Supporting or automating triage, e.g., suggesting questions, summarizing the conversation, or proposing an urgency level.

*B2. Self-care advice after triage* - Providing patients with self-care advice, or referring them to reliable sources, when a consultation is not needed.

*B3. Consultation preparation: patient-focused intake* - Helping patients structure their complaints before consultation through a digital intake and sharing the results with the professional.

**C.Clinical decision support**

*C1. Interpreting test and lab results -* Supporting healthcare professionals in interpreting blood test results and other diagnostic results (e.g., electrocardiograms, radiology reports).

*C2. Medication management and prescribing -* Supporting prescribing decisions (e.g., identifying interactions and contraindications) and periodic medication reviews.

*C3. Support with treatment plan and next steps -* Supporting the development of treatment plans and follow-up steps at the end of a consultation.

*C4. Support with differential diagnosis -* Assisting in generating a list of diagnoses before or during the consultation.

*C5. Support during consultations -* Providing real-time clinical support during consultations, such as suggesting follow-up questions or relevant physical examinations.

**D.Management of (chronic) diseases**

*D1. Identifying risks of (chronic) diseases -* Identifying patients at high risk of physical or mental health conditions from record data (e.g., to flag underdiagnosis or target preventive care).

*D2. Patient follow-up and monitoring -* Supporting patient follow-up and monitoring outside consultations, e.g., automated recall messages, medication reminders, or brief check-ins. *D3. Support for chronic care -* Supporting chronic care programs (e.g., diabetes, COPD), such as preparing check-ups or recording findings according to care protocols.

**E.Patient interaction and communication**

*E1. Clarification of medical information around consultation -* Explaining information around consultations and patient documents (e.g., specialist letters) in patient-friendly language.

*E2. Providing medical explanation and education -* Explaining general medical information not specific to the patient’s own care in accessible language (e.g. answering general health questions).

*E3. Improving professional patient communication -* Supporting professionals’ communication skills via consultation preparation, real-time suggestions, or feedback afterwards.

*E4. Self-management for diseases and lifestyle -* Supporting patients in behavioral change (e.g., smoking cessation, healthier eating) and chronic disease self-management.

SOAP: Subjective, Objective, Assessment, Plan; ICPC: International Classification of Primary Care; SNOMED-CT: Systematized Nomenclature of Medicine – Clinical Terms; COPD: Chronic Obstructive Pulmonary Disease.

## Results

Below, we present participant characteristics, the prioritization results, and the qualitative findings per application. Although prioritization concluded the interviews, we present it first to enhance readability.

### Baseline characteristics of participants

We included 31 participants, see Table 1 for their demographic characteristics.

**Table 1.** Demographic characteristics of all participants and by role.

|  |  | **All (n=31)** | **GP<sup>a</sup> (n=12)** | **AS<sup>b</sup> (n=7)** | **PN<sup>c</sup> (n=6)** | **PR<sup>d</sup> (n=6)** |
|---|---|---|---|---|---|---|
| **Age (years), median (range)** |  | 48.0 (23–81) | 44.0 (32–60) | 46.0 (23–55) | 44.5 (28–66) | 75.5 (56–81) |
| **Gender, female, n (%)<sup>e</sup>** |  | 18 (58) | 5 (42) | 7 (100) | 5 (83) | 1 (17) |
| **Experience (years), median (range)** |  | 18.0 (2–40) | 15.5 (2.5–25) | 18.0 (2–28) | 17.5 (7–40) | - |
| **LLM knowledge, median (range)<sup>f</sup>** |  | 4.0 (1–5) | 4.0 (2–5) | 3.0 (2–5) | 3.5 (1–5) | 3.5 (2–5) |
| **LLM use, n<sup>g</sup>** |  |  |  |  |  |  |
|  | *Daily* | 8 | 5 | 0 | 2 | 1 |
|  | *Weekly* | 13 | 5 | 4 | 2 | 2 |
|  | *Monthly* | 6 | 2 | 1 | 0 | 3 |
|  | *Sporadically* | 2 | 0 | 1 | 1 | 0 |
|  | *Never* | 2 | 0 | 1 | 1 | 0 |

<sup>a</sup>GP: general practitioner.
<sup>b</sup>AS: GP assistant.
<sup>c</sup>PN: practice nurse.
<sup>d</sup>PR: patient representative.
<sup>e</sup>Participants who did not identify as female identified as male or did not report their gender.
<sup>f</sup>Large language model (LLM) knowledge was self-reported on a scale from 1 (no knowledge at all) to 5 (highly knowledgeable).
<sup>g</sup>Large language model (LLM) use was self-reported as general frequency of use, combining personal and professional use.

### Prioritization of LLM support

Table 2 shows the ranking of applications by the number of high-priority selections (for which participants could select any number), with top three selections used to break ties. Some participants selected more than three top applications; all of these selections were included. Two applications were prioritized above all others: ‘Triage and urgency classification’ and ‘Summarizing consultations’ were both selected as high priority in 18 out of 31 interviews. Out of these two, ‘Summarizing consultations’ had the highest top 3 selection frequency (15 times). In third place overall, and the highest rank in the patient interaction and communication category was ‘Clarification of medical information around consultation’.

**Table 2.** Ranked prioritization of large language model applications overall and by role: number of high-priority votes (number of top 3 votesa).

| Application<sup>b</sup> | All<br>n=31 | GP<sup>c</sup><br>n=12 | AS<sup>d</sup><br>n=7 | PN<sup>e</sup><br>n=6 | PR<sup>f</sup><br>n=6 |
|---|---|---|---|---|---|
| *A1. Summarizing consultations* | **18 (15)** | 6 (5) | 6 (5) | 4 (4) | 2 (1) |
| *B1. Triage and urgency classification* | **18 (11)** | 8 (5) | 5 (4) | 1 (0) | 4 (2) |
| *E1. Clarification of medical information around consultation* | **14 (7)** | 3 (2) | 7 (3) | 2 (1) | 2 (1) |
| *C1. Interpreting test and lab results* | **13 (5)** | 6 (2) | 2 (1) | 2 (0) | 3 (2) |
| *D1. Identifying risks of (chronic) diseases* | **13 (5)** | 3 (1) | 3 (2) | 3 (1) | 4 (1) |
| *D2. Patient follow-up and monitoring* | **13 (4)** | 3 (0) | 4 (1) | 2 (1) | 4 (2) |
| *C2. Medication management and prescribing* | **13 (3)** | 3 (1) | 5 (2) | 3 (0) | 2 (0) |
| *C3. Support with treatment plan and next steps* | **12 (4)** | 7 (3) | 3 (1) | 1 (0) | 1 (0) |
| *D3. Support for chronic care* | **12 (4)** | 5 (2) | 2 (0) | 3 (1) | 2 (1) |
| *E2. Providing medical explanation and education* | **11 (6)** | 1 (0) | 4 (0) | 3 (3) | 3 (3) |
| *A2. Summarizing patient records* | **11 (5)** | 6 (2) | 1 (0) | 1 (1) | 3 (2) |
| *B2. Self-care advice after triage* | **11 (4)** | 4 (1) | 4 (1) | 1 (1) | 2 (1) |
| *C4. Support with differential diagnosis* | **10 (6)** | 6 (4) | 1 (0) | 1 (1) | 2 (1) |
| *A3. Processing medical letters* | **10 (6)** | 6 (3) | 2 (1) | 0 (0) | 2 (2) |
| *E3. Improving professional patient communication* | **10 (4)** | 3 (1) | 3 (1) | 1 (0) | 3 (2) |
| *A4. Writing referral letters* | **9 (3)** | 3 (1) | 2 (1) | 1 (1) | 3 (0) |
| *A5. Organizing inbox and correspondence* | **9 (2)** | 6 (2) | 1 (0) | 0 (0) | 2 (0) |
| *B3. Consultation preparation: patient-focused intake* | **8 (2)** | 3 (1) | 2 (0) | 0 (0) | 3 (1) |
| *A6. Coding of consultations* | **7 (2)** | 3 (1) | 1 (0) | 1 (1) | 2 (0) |
| *C5. Support during consultations* | **6 (2)** | 4 (1) | 1 (0) | 1 (1) | 0 (0) |
| *E4. Self-management for diseases and lifestyle* | **5 (1)** | 2 (0) | 0 (0) | 1 (1) | 2 (0) |

<sup>a</sup>Some participants selected more than 3 applications as their top priorities; all selections were included in the top 3 vote counts.
<sup>b</sup>The letter prefix indicates the application category (A: administrative tasks; B: pre-consultation and triage; C: clinical decision support; D: management of (chronic) diseases; E: patient interaction and communication). The number indicates the rank within the category.
<sup>c</sup>GP: general practitioner.
<sup>d</sup>AS: GP assistant.
<sup>e</sup>PN: practice nurse.
<sup>f</sup>PR: patient representative.

Table 3 shows the mean percentage of participants rating an application as high priority in each category, for all participants and by role. Overall, differences between categories were small (32%-41%). Preferences differed by role. GPs favored clinical decision support (43%), followed closely by administrative tasks and pre-consultation and triage (both 42%). Assistants favored pre-consultation and triage (52%), with patient interaction and communication a close second (50%). Practice nurses mainly favored management of (chronic) diseases (44%). Patient representatives also favored management of (chronic) diseases (56%), followed by pre-consultation and triage (50%).

**Table 3.** Mean percentage of participants rating an application as high priority, by category and role.^a^.

| **Category (no. of applications)** | **All (n=31) , %** | **GP<sup>b</sup> (n=12) , %** | **AS<sup>c</sup> (n=7), %** | **PN<sup>d</sup> (n=6) ,%** | **PR<sup>e</sup> (n=6) , %** |
|---|---|---|---|---|---|
| A. Administrative tasks (6) | 34 | 42 | 31 | 19 | 39 |
| B. Pre-consultation and triage (3) | 40 | 42 | **52** | 11 | 50 |
| C. Clinical decision support (5) | 35 | **43** | 34 | 27 | 27 |
| D. Management of (chronic) diseases (3) | **41** | 31 | 43 | **44** | **56** |
| E. Patient interaction and communication (4) | 32 | 19 | 50 | 29 | 42 |

<sup>a</sup>Values show the number of high-priority votes, normalized by the number of applications and participants, per category and role. Bold indicates the highest value per column.
<sup>b</sup>GP: general practitioner.
<sup>c</sup>AS: GP assistant.
<sup>d</sup>PN: practice nurse.
<sup>e</sup>PR: patient representative.

### Application of LLM support

For each application, we summarized the main benefits, risks, and requirements for its adoption identified by participants. Where participants suggested multiple functionalities within an application, we report these separately if their benefits, risks, and requirements differed. The full coding overview is provided in Supplementary File 3.

## A. Administrative tasks

### A.1. Summarizing consultations

The primary benefits of LLMs for summarizing consultations were workload reduction and more attention for the patient during consultations. Professionals felt that summaries by LLMs were more complete and consistent than manually written notes, improving information transfer among professionals.

However, LLM use could negatively affect workflows. Automated summarization may eliminate the clinical reasoning that occurs during manual note writing. Adopting the new workflow would require verbalizing findings and the physical examination and manually copy-pasting summaries into the EHR when direct integration is unavailable.

Data safety and privacy were also raised as concerns. Patient representatives emphasized that explicit patient permission for recording was required, although explaining the tool to patients could be time-consuming.

Participants also raised concerns about the quality of the summaries. Quality could be negatively affected by inclusion of irrelevant non-consultation information, omission of important non-verbal cues, or hallucinations. To mitigate these risks, participants requested automatic filtering of irrelevant information, support for multiple languages and dialects, and model training on real patient consultations. Human validation was seen as a crucial safeguard for summary quality.

### A.2. Summarizing patient records

Benefits of LLM support for summarizing patient records were time reduction for professionals and easier information transfer to third parties such as insurers. Participants, mostly GPs, thought that a better patient overview would lead to more informed medical decisions, improving quality of care. This was seen as particularly helpful for locum or substitute professionals.

However, for regular GPs with their own patient population, summaries were seen as less valuable. Moreover, as current EHRs already provide a patient history through coded episodes, some participants raised doubts about the added value of LLM support. Participants also raised concerns about hallucinations in LLM generated summaries.

Functional requirements were mostly addressed by GPs. They required summaries to be fast and have an adjustable period (e.g., the last x months). The application should retrieve information from multiple sources, including legacy data, medical letters, and external hospital data. GPs also envisioned combining it with speech-to-text software to enable real-time patient information retrieval during consultations.

### A.3. Processing medical letters

The primary benefit of LLM support for processing medical letters was time reduction, predominantly raised by GPs. Professionals also anticipated missing fewer follow-up actions from incoming letters. One GP primarily valued this application as groundwork for future digitalization efforts that rely on specialist information, such as patient record summarization (*Application A*.*2*). However, several participants questioned the added value of LLMs, as specialist letters often already include a conclusion, and some practices store letters in full regardless.

Participants raised two main concerns: missing important correspondence and reduced engagement with patients due to automation. To mitigate these risks, they suggested an alert system for important letters combined with human validation. Professionals further noted standardization as a practical challenge, as each practice handles correspondence differently. To accommodate this, they requested customizable options such as adjustable summary length.

### A.4. Writing referral letters

Participants primarily valued LLMs for making referral letter writing more efficient. LLMs could also ease the overall referral process, for example by helping select the right care provider. However, participants questioned what LLMs would add beyond the digital referral platform already used in the Netherlands. Some would welcome LLM-based extensions of this platform, while others noted that such improvements would not necessarily require LLMs.

Views on quality were mixed. Some professionals expected LLMs to improve referral letter quality, others questioned whether an LLM could accurately capture the referral question. One practice nurse noted that LLMs cannot capture the patient’s subjective context, which is important in some referrals, such as those to mental health care.

To address these concerns, participants outlined several requirements. The GP should retain responsibility for the referral reason, with the LLM handling remaining content. EHR integration was considered essential for retrieving relevant patient information. Generated letters should be reviewed by the professional. Patient representatives proposed an additional patient review step.

### A.5. Organizing inbox and correspondence

GPs and assistants, who handle most correspondence, expected time savings as the main benefit. This was particularly relevant for processing overloaded inboxes during busy periods, such as after weekends and holidays. They also anticipated more timely care, as it could prevent delays in handling urgent messages.

However, professionals questioned the added value for three reasons. First, time savings were seen as limited, particularly for smaller practices. Second, all patient information must be processed in the EHR regardless, limiting efficiency gain. Third, existing EHR functionality already supports classification of urgent messages. Professionals raised two additional implementation concerns: the difficulty of standardizing across practices and the risk of urgency misclassification by the model.

As a functional requirement, one GP suggested the automatic scheduling of follow-up tasks arising from incoming correspondence, such as booking lab tests requested in a specialist letter.

### A.6. Coding of consultations

Most insights into coding were addressed by GPs, who hold the main responsibility for this application. Within this application, several subtasks emerged. First, LLMs could assist with linking uncoupled consultation notes or episodes, reducing backlog. This was seen as particularly useful when migrating patient data from another practice. Second, LLMs could support coding of incoming medical letters or new consultations, reducing manual labor. Third, LLMs could support re-coding, correcting wrongly coded entries. Fourth, LLMs could group related codes together, reducing redundant or overlapping codes and improving professionals’ overview of patient information. Across these subtasks, the primary benefits were improved EHR coding quality and reduced time spent on coding.

However, participants raised several concerns about added value. Many felt that coding is currently not time-consuming, or that existing support is already sufficient. Participants questioned whether algorithmic approaches other than LLMs might be better suited, noting that current LLM coding performance is still lagging. As a result, human validation would remain necessary, potentially limiting the overall efficiency gain.

Some GPs also envisioned extending the current coding methodology, which focuses on diagnosis only. They suggested using an LLM to automatically code the full consultation content, including medical history and physical examination, capturing clinically relevant but currently unstructured information. This could yield richer and more searchable patient data, supporting applications such as *summarizing patient records* (application A2). These codes could also enable more retrospective and prospective clinical analysis, such as *identifying risks for chronic diseases* (application D1) However, participants raised concerns about limited transparency and the risk of unintended recording of confidential information. Participants noted that current EHR functionality would need to be extended to support and use this extended coding system.

## B. Pre-consultation and Triage

### B1. Triage and urgency classification

Participants distinguished two forms of LLM support for triage: one facing the assistant and one facing the patient.

For assistant-facing support, two subtasks emerged, both relying on speech-to-text transcription of the triage call. First, LLMs could reduce administrative workload by automating triage call documentation, similar to *summarizing consultations* (application A1). Second, LLMs could improve triage quality through real-time conversation support, for example by suggesting triage questions or urgency levels. Several requirements were raised. The application should connect to telephone services and integrate both patient data and national triage guidelines. It should be able to handle unstructured conversations, such as patients presenting multiple complaints, and offering multilingual support. Where needed, assistants should be able to override LLM’s recommendations, for instance to adjust urgency level. The main concerns were that this support would not resolve the assistant’s shortage and might yield only limited time savings.

Patient-facing support was seen as having a greater impact on work demand. Suggested workflows were a dynamic triage chatbot or questionnaire completed before contacting the practice. Through increased adoption of digital triage, participants envisioned fewer patient contacts, shorter waiting times, and improved accessibility. This could reduce the assistant staffing shortage. However, several strong concerns were raised. First, LLMs lack human intuition and “gut feeling” critical to triage quality. Second, patient resistance could limit adoption. Third, digital tools may reduce accessibility for patients with low digital literacy or socioeconomic status. Fourth, GPs highlighted clinical safety risks, foreseeing accuracy limitations and misjudgment of urgency. For safe implementation, professionals suggested routing difficult or urgent cases to assistants through traditional channels such as the phone. Another suggestion was limiting the tool to structuring the patient’s question without giving advice, leaving completed cases for a GP or assistant to process.

Overall, participants favored a cautious, gradual roll-out rather than rapid adoption. They preferred LLMs to first support assistants before taking on more autonomous triage. They also stressed that any patient-facing tool should follow the triage process assistants already use by phone rather than introduce a new logic.

### B2. Self-care advice after triage

Patients can use public self-care services before contacting the GP. Participants wanted more patient requests to be handled through these services to reduce demand in the practice, but were not specific about how LLMs could contribute. Assistants also give self-care advice by phone after triage, often drawing on the same public sources. An LLM connected to the phone line could generate this advice automatically, tailoring information from public self-care sources to the patient using triage conversation and patient data. Some participants felt this could make giving self-care advice more efficient, improve its quality, and make it more evidence-based and consistent across providers. However, one GP questioned the added value over simply referring to the public self-care sources. Participants warned that the LLM might cite incorrect sources, so it should draw only on high-quality self-care information. The generated advice should also be validated by a professional before it is sent to the patient.

Several concerns applied to digital self-care in general. Patient representatives noted that limited digital literacy could restrict patient adoption. To improve accessibility, GPs suggested offering multiple communication channels, such as spoken rather than written text. GPs also raised the financial consequences of non-billable self-care pathways and uncertain legal responsibility.

### B3. Consultation preparation: patient-focused intake

Participants noted that LLMs could improve patient preparedness by helping patients structure their care question, understand care options, and provide a more complete pre-consultation history. This could support patient autonomy and shared decision making. In addition, it could lead to shorter consultations by reducing time spent on medical history. Professionals also foresaw fewer consultations due to earlier redirection of unsuitable care questions and fewer follow-up visits because of missing preparatory information.

However, professionals addressed two potential negative impacts on consultations: the risk of diagnostic tunnel vision and asking patients the same question twice. Automating part of medical history was also seen as potentially reducing job satisfaction. GPs proposed several functional requirements: EHR integration, combining consult preparation data with triage information, and GDPR-compliant data transfer from patient to practice.

Participants also raised several patient-related barriers. Adoption could be limited by resistance to pre-consultation instructions or limited digital literacy. Participants also questioned whether all patients could accurately report their complaints digitally. Offering multiple input formats, such as speech, was suggested to mitigate these risks.

## C. Clinical decision support

### C1. Interpreting test and lab results

LLM support for interpreting test and lab results was valued by GPs for its potential to improve clinical decision-making. It could support them by expanding clinical knowledge and identifying appropriate follow-up actions. GPs also saw potential to improve patients’ understanding of diagnostic results through patient-friendly messages. Overall, most participants anticipated a reduction in workload through LLM support.

However, GPs questioned the added value of LLM support, as routine test interpretation does not require specialized AI assistance. Some GPs viewed LLM involvement as a threat to clinical autonomy, potentially further limiting adoption. Additional concerns included overreliance on automated interpretations and ambiguity about legal responsibility.

Professionals highlighted specific risks around quality. Low quality or missing patient data and defensive model behavior could degrade results. Patient-specific nuances and edge cases near cut-off values were seen as particularly hard for LLMs to interpret. Human validation was seen as essential by all participants.

Key input data requirements included clear cut-off values, the clinical indication for the test, patient context, and historical results. Professionals preferred integration of LLM support directly within the EHR inbox, where results are received.

### C2. Medication management and prescribing

Professionals valued deprescribing support to improve medication oversight and to reduce unnecessary medication use. They also suggested support for polypharmacy and medication reviews in collaboration with pharmacy. LLMs could reduce the time these require or increase the capacity to carry them out.

Professionals said that prescription support by LLMs could improve overall quality of prescribing by preventing dosing errors and signaling missed treatment opportunities. They also expected time savings. Suggested capabilities included medication conversion and substitution support, interaction alerts, repeat-prescription automation, health insurance reimbursement checks, and medication misuse monitoring (e.g. for benzodiazepines). However, they also questioned the added value of LLMs, as current EHRs already provide built-in prescription assistance.

Participants emphasized that LLM prescription support should be based on up-to-date medical guideline information. Another suggested prerequisite was the availability of an up-to-date medication overview. However, participants noted that this is difficult to maintain, as medication information is not always shared consistently between healthcare providers.

One GP was skeptical about an LLM’s ability to give accurate medication suggestions using its internal knowledge. Instead, a tool-calling, agent-based system was suggested, in which the LLM uses external structured medicine information to create recommendations.

### C3. Support with treatment plan and next steps

LLMs could support treatment planning in several ways. Participants primarily valued more efficient guidelines and protocol searching, and the flagging of blind spots or missed actions. GPs thought this could reduce specialist referrals, as they could resolve more questions themselves. Doctor assistants similarly expected to become more self-sufficient, relying less on the GP. Extending this application, professionals suggested automating follow-up steps, such as ordering labs or enrolling patients in external (self-)care programs.

Several concerns were raised. One GP worried that increased screen use might annoy patients and suggested being transparent about using an LLM support system. One practice nurse stressed that patient-specific context is crucial, making LLM suggested treatment plans hard to standardize. Another practice nurse feared deskilling of new GPs who become overly dependent on the system’s clinical support. To guard against this and preserve the human aspect of care, several participants emphasized the importance of clinician override and autonomy.

### C4. Support with differential diagnosis

Participants thought LLMs could support critical thinking for differential diagnosis and difficult cases. GPs and assistants expected improved diagnostic accuracy. Professionals also noted that differential diagnosis support could serve as the foundation for other applications, such as s*upport with treatment plan and next steps* (task C3).

However, differential support could disrupt the GP workflow. One GP noted that general practice constantly involves decision-making under uncertainty, limiting the value of detailed suggestions. Others feared constant self-doubt, and that reviewing suggestions would add work. To contain this, a doctor assistant suggested limiting the diagnostic scope rather than overloading GPs with possibilities.

Overreliance was another concern, making clinician override and autonomy an important prerequisite. Individual GPs also questioned the added value of LLMs over existing algorithms and raised the variability of LLM output as a concern. For successful implementation, they stressed that both patient information and guidelines should be integrated.

### C5. Support during consultations

Professionals valued live consultation support for offering critical-thinking assistance and reducing clinician blind spots. They also thought it could improve history-taking, for example by prompting clinicians to ask about specific alarm symptoms. One GP added that LLMs could suggest or automate standard question formats, such as risk-score frameworks for conditions like gout. Another GP felt it could support professional development and job satisfaction, but doubted colleagues more resistant to change would accept it at all.

The main concern was that it could draw attention away from the patient, although one GP imagined it reshaping consultation style: by reviewing LLM recommendations together with the patient, it could strengthen shared decision-making. This would, of course, require openness with patients about LLM use. For the system to work well, GPs noted it would need integration of the patient record, differential diagnoses, and treatment guidelines. Overall, participants stressed that clinicians should always be able to override or ignore recommendations.

## D. Management of (chronic) diseases

### D1. Identifying risks of (chronic) diseases

Professionals felt LLMs could reduce workload in preventive care, for example by automating screening such as drafting patient lists of risk groups. More broadly, it could expand preventive care capacity, currently limited by time constraints. Participants felt it could improve care quality by detecting overlooked patients or enabling more timely care.

The main undesired consequence, raised solely by GPs, was that it could create more work for the practice. One practice nurse stressed that the practice would need more budget to finance additional care. Another GP said the tool should not be too broad but focused on specific care paths. GPs described two contrasting workflows: one in which the tool runs periodically outside the consultation, and another during the consultation. The first GP argued it should not disrupt consultation flow, while the second argued that placing it outside the consultation would lead to preventive care being overlooked.

Some professionals questioned the added value of LLMs over current risk-screening tools or questionnaires. Participants noted this is not inherently an LLM application, and that rule-based algorithms might be better suited. They added that using LLMs here would require very high-quality, granular patient data to enable text-based risk screening.

### D2. Patient follow-up and monitoring

Within this application, three main application areas emerged. First, LLMs could support follow-up and action reminders. Participants envisioned a chatbot checking in with patients after consultations. For example, reminding them of follow-up actions or treatment. Patient representatives thought this might improve medication management and adherence. Doctor assistants thought it could increase patients’ sense of being cared for. The main risks raised by both professionals and patients concerned patient interaction: low digital literacy, low engagement, and the risk of overburdening could all limit adoption. To mitigate these, participants suggested personalization, patient-friendly design, and the option to opt out. Professionals also proposed a task-scheduling function to reduce missed follow-up actions on their side.

Second, recalling and managing chronic patients was a frequently raised challenge. Professionals felt LLMs could address this by automating the (re)calling process. Many highlighted time savings, alongside reducing patients lost to follow-up. One GP noted, however, that simple digitalization without an LLM would suffice here. A further concern was loss of patient oversight through automation. To mitigate this, professionals suggested an alert system for patients unresponsive to recalls. They added that an online recalling system should be reserved for patients with sufficient digital skills, while other patients should be called directly.

Third, LLMs could support patient self-measurement systems by analyzing and filtering measurements. Patients felt this could make sharing information with the practice easier. Professionals thought it could reduce live consultations for GPs and practice nurses. However, GPs and patients were concerned it would be hard to fit into the current workflow and could increase workload. They also thought patient adoption might be low. For implementation, data should flow automatically into the EHR, possibly via wearables. To manage volume, participants proposed that the system should flag only measurements requiring attention. One GP noted that self-monitoring should only be available to stable patients, and that personal contact should be preserved.

### D3. Support for chronic care

Because chronic care is heavily protocolized, LLMs could enable more digital care by automating (part of) a consultation. Participants felt that this could reduce overall workload and free up consultation time for relational care. However, professionals feared this time saving might be redirected to other tasks rather than personal contact, leaving patients with increasingly digital care. They were concerned about a weakened patient-provider connection and the loss of personal non-medical information. They also noted that digital chronic care might lower patients’ motivation for treatment and lifestyle adherence. Moreover, limited digital literacy among chronic patients could limit adoption of digital care pathways.

Relying on speech-to-text technology, LLMs could support professionals, especially practice-nurses, during or after chronic care consultations. Related to *consultation summarization* (application A1), professionals suggested automating the recording of standard measurements required by chronic care protocols. Similar to *support during consultations* (application C5), LLMs could offer care suggestions based on chronic care guidelines, reducing search time and improving guideline adherence. One GP noted that patient information should be incorporated to allow personalized recommendations. However, practice nurses raised two concerns. First, individual patient complexity would limit such automated support. Second, protocols are already visible in the EHR, reducing the added value of an LLM.

## E. Patient interaction and communication

### E1. Clarification of medical information around consultation

Both professionals and patients felt LLMs could make medical information more understandable for patients. Mentioned information sources included specialist letters, patient files, and consultations. Patient-friendly consultation summaries were suggested most often. Overall, they felt this support could improve the patient–provider relationship and reduce workload through fewer follow-up calls and visits. GPs also envisioned LLM support for answering e-consultation messages, saving time while improving answer quality. Beyond patient support, assistants mentioned direct benefits for themselves, for example in translating complex specialist letters into more understandable language.

Professionals particularly valued support for vulnerable groups such as low-literate patients, the elderly, non-native speakers, or those hesitant to ask questions during consultations. At the same time, they worried that limited digital skills could hinder adoption among these patient groups. To mitigate this, professionals and patients suggested multiple information formats (e.g. speech) and delivery channels (e.g. text message, email, patient portal). They also valued multilingual support. GPs were divided on responsibility for the tool: some felt it should lie with the GP, others with the patient. If the GP is responsible, one noted that patient-friendly summaries would need GP review, doubling the validation burden alongside the professional consultation summary. Under a patient-led approach, another GP worried that two parallel patient files could emerge (one for the patient, one for the GP), doubling information sources.

### E2. Providing medical explanation and education

Participants discussed using LLMs to provide medical information through a general health chatbot interface. They felt this could improve patient preparation, self-care, and self-management before consultations. Participants emphasized it could raise the overall quality of medical information patients encounter, by offering an alternative to unvalidated online sources. One GP valued that LLMs could provide reliable information in real time, as patients often want immediate answers at any time of day or week. Professionals also thought LLMs could support patients after consultations by providing general health information, for example about their diagnosis. This was seen as especially useful for onboarding new chronic patients. Overall, professionals envisioned this support saving time through reduced patient contact.

Participants raised several patient-related concerns. Adoption might be limited by low digital literacy, doubts about credibility, preference for human explanation, or mismatched language level. As requirements, patient representatives suggested adjustable language level, references to sources, high-quality information inputs, multiple formats (e.g. speech or visual), and multilingual capabilities.

Views on organizing the tool in practice were mixed. Some professionals preferred a central initiative outside GP workflow, while others wanted to retain control themselves. Some GPs were concerned about unclear ownership. They feared that promoting a public tool would lead to patients interpreting its answers as direct advice from them, or that an external tool might use their practice label without their oversight.

### E3. Improving professional patient communication

Participants thought LLMs could improve professional-patient communication before, during, and after consultations. This support could be generic, or patient-specific by drawing on a live consultation transcript. Before consultations, participants noted LLMs could help prepare personalized communication strategies and support with difficult conversations. During consultations, the most frequently raised use case was support with motivational interviewing. After consultations, participants valued LLM-supported feedback, for example by analyzing difficult conversations or questioning style. They appreciated that such feedback was available on demand and preferred it over self-review or peer-review because it felt less confrontational. Participants noted this feedback could add value in educational settings for junior staff, as well as for senior staff through recurrent performance evaluation.

However, participants were uncertain whether LLMs could adequately assess communication quality. They felt LLMs could not detect subtle cues or could overlook situational context. Participants also anticipated reluctance among professionals to adopt an LLM-powered feedback system, especially during consultations. To increase adoption, one GP suggested linking the tool to accreditation credits as a form of post-qualification training.

### E4. Self-management for diseases and lifestyle

Both practice nurses and GPs felt that LLMs could improve existing self-management programs, through increased personalization and accessibility based on patient-specific information. Professionals thought that improved self-management programs could increase patient autonomy and lifestyle awareness, freeing up clinician time for more urgent patient care.

However, assistants and patient representatives were concerned about low patient engagement and the digital literacy of target groups such as chronic patients. They also raised safety risks for serious issues such as mental health, though one GP felt these tools could be a safer alternative to unsupervised commercial platforms.

Participants were divided on how such a tool should be organized. Most preferred a central initiative outside the GP workflow, while one GP favored a tool linked directly to the practice to increase patient engagement.

## Discussion

### Prioritization of LLM support

This interview study offers insights into 21 LLM applications across 5 categories in Dutch general practice. Through structured prioritization, we identified promising candidates for further development. The two highest-priority applications were summarizing consultations and triage support. Both were also prioritized in earlier research [24, 25] and are already widely adopted in practice [28, 29]. However, current digital triage applications are mostly rule-based or use traditional machine learning [30]. LLMs could enable new applications, such as conversational triage chatbots or real-time decision support during triage calls. The third most prioritized application, clarification of medical information around the consultation, was not prioritized in prior research. Participants particularly valued a patient-friendly summary of the consultation, extending automated notetaking from professionals to patients.

Our study extends prior LLM prioritization research [25] by capturing the priorities of practice nurses, GP assistants, and patient representatives alongside those of GPs. Although interest was shared across categories and stakeholder groups, participants often prioritized applications tied to their own role (e.g., assistants favored triage). Both practice nurses and patient representatives prioritized chronic disease management, reflecting a need to meet rising chronic care workload and patient demand [31, 32]. Among GPs, clinical decision support received high priority. This differs from an earlier survey, in which fewer GPs considered LLMs useful for clinical applications (such as differential diagnosis) than for non-clinical applications (such as consultation notes) [25]. Overall, our prioritization shows that interest in LLMs spans multiple areas of general practice, in line with their reported potential [14].

### Application of LLM support

Although prioritization showed broad interest in LLMs, our qualitative analysis revealed three important considerations for their implementation: whether an LLM is needed, whether it relieves workload, and how its benefits weigh against its risks.

#### Assessing the need for LLMs

Not every application requires an LLM, and several factors influence whether an LLM is needed. First, the added value of an LLM differs by application: some workflows are already efficient and cannot be optimized much further. For example, participants said that consultation coding is currently not very time-consuming, so LLM support would add little value. Second, LLMs are not always the right technology: simpler or deterministic digital tools could support many workflows equally well. Participants frequently raised recalling and managing chronic patients as a challenge, yet this could be addressed through digitalization without LLMs. Third, existing systems may already suffice. For example, in the Netherlands, there is already a platform that streamlines referrals between providers, reducing the need for LLM-written referral letters. Taken together, these factors suggest that applying LLMs too broadly risks creating overly complex or unnecessary digital solutions. LLMs should therefore be treated as a possible solution, not a universal remedy. This view aligns with earlier findings that primary care stakeholders see AI as “just another tool in the toolbox”, valued for its contribution rather than the technology itself [18]. Our application-level analysis offers a starting point for identifying where LLMs are the right tool.

#### Establishing workload relief

If an LLM is the right tool, the main benefit participants expected was workload reduction. However, participants also pointed out that an LLM intervention may generate work, thereby limiting these gains. This aligns with previous research suggesting that digital health interventions can increase a clinicians’ burden rather than relieve it [33-35]. The concern was raised especially for clinical support applications, such as help with management plans or differential diagnosis, which may produce additional suggestions the professional has to review or act on. Patient-facing applications, such as a tool for sharing self-measurements with the practice, may similarly raise care demand or be difficult to fit into practice workflows. Establishing whether an LLM actually relieves workload is an essential step before implementation.

#### Weighing benefits against risks

Even when an LLM is needed and relieves workload, its implementation carries risks that need to be weighed against the benefits. We highlight three main risks raised by participants.

The first risk is mistakes. LLMs are well known to hallucinate [36]. A human validation step may be needed to safeguard quality; for example, with ambient scribes, clinicians routinely revise the generated summaries to correct inaccuracies [37]. This human check costs time and thus limits the potential time savings of LLMs. Moreover, it is unclear what level or type of errors are acceptable in the first place. Medical professionals also make mistakes, yet it is not obvious that we should tolerate the same from an LLM [38]. Application design should consider the level and type of errors we are willing to correct or accept.

The second risk is reduced personal care and accessibility. Participants repeatedly stressed that digitalization through LLMs might reduce personal care. Personal care is an important driver of care quality and patient satisfaction in general practice [39]. Although participants noted that the freed-up time could be redirected to relational care, it is likely that the time will be used for other tasks given the high overall workload. Related, participants frequently mentioned decreased accessibility as a risk. Because the accessibility of digital tools in general practices depends on patient characteristics such as health and digital literacy [40], increased digitalization might exclude vulnerable patients groups. Application design should consider how to preserve personal contact and accessibility while retaining the benefits of digitalization.

The third risk is negative effects on professionals. In line with previous research, participants mentioned overreliance [41], automation bias [41], and deskilling [42] as concerns. Participants also noted that LLM support may conflict with core values of general practice, such as decision-making under uncertainty and gatekeeping. LLM might add uncertainty, create false certainty, or misalign with the values of the treating physician or healthcare context [43]. Application design should consider how to align with the professional autonomy, skills, and values of general practice professionals.

### Implications

The risks of LLMs call for design choices, which interact with each other and with the expected benefits. For instance, assistant-led triage with LLM suggestions may reduce mistakes and retains personal contact, but limits time savings and may invite automation bias. By contrast, a fully automated chatbot maximizes efficiency but risks undetected errors, less personal contact, and lower accessibility. Rather than pointing to a single best design, these choices form a set of trade-offs that must be weighed. The design should be selected through co-creation with professionals and patients, who can judge which trade-offs are acceptable.

The relevant stakeholders depend on the application. Patient-facing tools should involve the vulnerable patient groups most at risk of being disadvantaged by digital technology. Broader implementation questions may require input from stakeholders beyond our sample, such as EHR vendors, regulators, and insurers. Finally, validation studies typically evaluate a single design. We suggest instead comparing multiple design variants within a single validation study, so that both the workload and the risk trade-offs identified here can be analyzed directly.

### Strengths and Limitations

The strengths of our study are threefold. First, we introduce a new overview of LLM applications covering a substantial portion of general practice workflow, grounded in literature and validated with participants. Second, we included multiple stakeholder groups, adding the perspectives of patients and non-GP professionals. Third, we conducted an in-depth application-level analysis to shape the priorities, going beyond a ranked list or higher-level thematic analysis.

However, some limitations should be addressed. First, our sample may be subject to selection bias. We did not purposively sample for diversity in LLM use or professional experience. Although we included participants both with and without LLM experience, most showed an affinity for the topic. This was to some extent necessary, since envisioning future applications requires some familiarity, but it may have made the insights less critical. Second, participants reasoned about applications they had not used. Our findings therefore reflect expectations rather than evaluations. As a result, harder-to-imagine applications may have been undervalued. Moreover, as LLM tools develop rapidly, priorities may shift as applications mature or become adopted in practice. Third, although our fixed application list enabled structured comparison of prioritization and qualitative insights, we may have missed other high-impact applications. We mitigated this by validating the list with professionals beforehand. Fourth, the sample was small for a quantitative prioritization, particularly within stakeholder groups (n=6–12). Our prioritization results are therefore descriptive and sensitive to individual votes and should be interpreted alongside the qualitative findings.

### Conclusions

GPs, assistants, practice nurses and patient representatives showed interest in LLM support across many areas of general practice. However, realizing LLM support requires three steps. First, the need for the LLM should be critically assessed against other alternatives that may be better suited. Second, it should be established whether the LLM actually relieves workload. Third, the application should be designed so its benefits outweigh the risks of mistakes, reduced personal care and accessibility, and negative effects on professionals. Our application-level analysis and prioritization offer a starting point for this assessment and further design.

## Supporting information

Supplementary File 1 - Interview Protocol

Supplementary File 2 - Interview Guide

Supplementary File 3 - Code System

## Data Availability

All data produced in the present study are available upon reasonable request to the authors.

