## Supplementary File 1 - Interview Protocol for "Applications of large language model in general practice: a qualitative interview study with professionals and patients"

### Introduction (5 minutes)

- Introduce the researcher, the project, and the purpose of the interview:
  - Self-introduction (name, role, and institutional affiliation)
  - Overview of the research project
  - Purpose of the interview
- Explain the structure of the interview:
  - **Part 1**: Review a structured list of applications for which LLMs may support general practice staff and patients. This list was compiled from the literature and prior stakeholder input.
  - **Part 2**: Prioritize the applications the participant considers most relevant for LLM support.
- Confirm informed consent, including consent for audio recording and the use of pseudonymized data.
- Start the recording.
- Ask the participant to introduce themselves:
  - *Professionals*: current role, relevant experience in general practice, and familiarity with LLMs.
  - *Patient representatives*: background in patient representation and familiarity with LLMs.

### Part 1: Application Analysis (30–40 minutes)

The application analysis follows the same core structure for all participants, with minor adjustments depending on their background. The aim is to learn which applications participants support, which they do not, and why.

#### Introduction (~3 minutes)

- Briefly explain what large language models (LLMs) are and give relevant examples of their potential use in general practice.
- Tailor the depth of the explanation to the participant’s familiarity with LLMs.

#### Eliciting unprompted ideas (~2 minutes) - *professionals only*

- Ask whether the participant already uses LLMs in their work on a regular basis.
- Ask the participant to name potential LLM applications beyond those they already use, i.e., tasks they think could benefit from LLM support. This question is optional; participants may skip it if no relevant tasks come to mind.

#### Explaining the applications and categories (~3 minutes)

- Introduce the application list, explaining that it is based on a literature review and initial stakeholder input and that the applications are grouped into categories.
- Give a brief overview of the categories so that the participant understands the types of applications covered.
- Explain that the next part of the interview will go through the predefined applications category by category.
- Encourage the participant to ask for clarification whenever anything is unclear.

#### Category-by-Category Review (~25–30 minutes)

- For each application category:
  - Introduce the category.
  - Present and explain each application within the category.
  - Ask the participant which applications in the category are and are not suitable for LLM support, and why.

### Part 2: Prioritization (10 minutes)

After the application review, participants are asked which applications they consider most important for LLM support.

- Present the full application list to the participant.
- *Prioritization step 1 – Identify high-priority applications:* Ask the participant to select the applications they consider the highest priority for LLM support (no fixed number) and to explain their choices.
- *Prioritization step 2 – Rank the top applications:* From the high-priority applications, ask the participant to select their personal top three, rank them in order of importance, and explain the ranking.

### Closing (2–3 minutes)

- Thank the participant for their time and insights, and briefly explain how their input will contribute to the research.
- Offer to share a summary of the study results once available, if the participant is interested.
- Inform the participant about opportunities for further involvement, such as future co-creation sessions or validation workshops.
- Invite any final reflections or suggestions, particularly on the future implementation of LLMs in general practice or directions for further research.
