## Supplementary File 2 - Interview Guide for "Applications of large language model in general practice: a qualitative interview study with professionals and patients"

1. **Introduction**

| **Topic** | **Script** | **Checklist / Notes** |
| --- | --- | --- |
| Explanation of interview | Prompt 1A |  |
| Interview structure | Prompt 1B |  |
| Consent form | Prompt 1C | □ Consent form signed |
| Recording | Prompt 1D | □ Recording explained  □ Recording started |
| Self-introduction | For professionals:   - What is your experience and current role within general practice? - How long have you worked in this position and in similar positions? - How would you rate your familiarity with large language models (such as ChatGPT) on a scale from 1 (not familiar at all) to 5 (very familiar)? - How often do you use language models such as ChatGPT (daily/weekly/monthly/yearly/never)?   For patient representatives:   - Could you briefly describe your experience as a patient representative? (formally within an organization or in a personal capacity) - How would you rate your familiarity with large language models (such as ChatGPT) on a scale from 1 (not familiar at all) to 5 (very familiar)? - How often do you use language models such as ChatGPT (daily/weekly/monthly/yearly/never)? | Years of experience in general practice:  Familiarity with language models (1-5):  Use of language models: |

2. **Task validation**

| **Topic** | **Script** | **Checklist / Notes** |
| --- | --- | --- |
| Introduction to language models | Not familiar with language models: Prompt 2A1  Somewhat familiar with language models: Prompt 2A2  Very familiar with language models: Skip |  |
| Open exploration *(professionals only)* | - Do you already use models such as ChatGPT in your work? - Can you name one or more tasks in your work that you think could be supported by a language model such as ChatGPT? *(optional)* This can be anything, for example tasks related to administration, communication with patients, or summarizing information.   - How do you think language models could help with these tasks?   - Why would support with these tasks be useful or necessary? |  |
| Explanation of categories and tasks | Prompt 2B |  |
| Discussion of tasks per category | - Are there tasks within this category that you would prefer not to be supported by a language model/AI/ChatGPT?   - Why not? - Are there tasks within this category that you think are not very relevant or worthwhile to support with AI? (e.g., because they take little time or already go well without technology)   - Why not? |  |

3. **Task prioritization**

| **Topic** | **Script** | **Checklist / Notes** |
| --- | --- | --- |
| Prioritization – step 1 | - Which tasks on this list would you most like to have supported by a language model/AI/ChatGPT?   - Why do you consider these tasks important?   - What makes the other tasks less important to you? |  |
| Prioritization – step 2 | - Of the tasks you marked as important, which three are the most important to you?   - Why do you consider these three tasks the most important?   - What makes the remaining tasks less important? - Could you rank these three from most to least important?   - What is your reason for this order? |  |

4. **Closing**

| **Topic** | **Script** | **Checklist / Notes** |
| --- | --- | --- |
| Closing | Prompt 4A |  |
| Staying informed | - Would you like to be kept informed of the results of this study? | Yes / No |
| Future involvement | - Would you like to take part later in the project in a workshop or other activity to further develop possible AI solutions?   - If so, may we record your contact details for this purpose? | Yes / No    Yes / No |
| Closing questions | - Do you have any ideas or points of attention for the next steps of this study? - Is there anything else you would like to add that we have not discussed today? |  |

**Example prompts**

**Prompt 1A – Explanation of interview**

I am a researcher at [*describe involved institutes].* We are investigating how AI models such as ChatGPT can be used in general practice care. We want to map out which tasks AI could potentially help with, and how professionals in general practice and patients feel about this. We are interested both in tasks where you see opportunities and in tasks for which you think an AI model is specifically not suitable. We are investigating this by means of interviews. We will publish the results in a scientific article, so that other researchers and companies can also take the views of those directly involved into account in future AI applications in general practice care.

**Prompt 1B – Interview structure**

The interview consists of two parts:

a. In the first part, we will discuss a number of pre-prepared tasks for which we think AI models could support general practice care. We will explain these tasks using several categories. We would like to hear whether you have any questions or comments about them, which tasks you find valuable, and which tasks you think should specifically not be performed by AI.
b. Then, in the second part, we will try together to prioritize these tasks: which do you consider most important, and why?

**Prompt 1C – Consent form**

You received a consent form in advance. Do you have any questions about it?

- *If signed*: Thank you for signing and returning it.
- *If not signed*: Before we can start, we need to ask you to sign this form.

**Prompt 1D – Recording**

With your permission, we would like to record the interview so that we can properly transcribe and analyze it later. Your name will not be used in the analysis; we will link your data to an anonymous code. Do you agree to the recording? If so, I will start it now.

**Prompt 2A1 – Introduction to language models (not familiar)**

A language model is a computer program that has been trained to work with text. The best-known example is ChatGPT. Such a model can, for example, answer questions, give explanations, or summarize information. You can think of it as a kind of smart assistant you can chat with.

General practice involves a lot of work with text. Such a model could therefore potentially help with all kinds of tasks. For example: summarizing a conversation between a doctor and a patient, rewriting a medical letter in understandable language, or answering a medical question from a healthcare professional.

We would like to discuss with you for which tasks you think such a model could be useful, and for which it could not.

From now on, we will refer to these kinds of systems as "language model" or "AI model"; this includes ChatGPT.

**Prompt 2A2 – Introduction to language models (somewhat familiar)**

A language model is an AI system that has been trained on large amounts of text. This enables it to analyze texts and generate new texts. It can, for example, answer questions, give explanations, or summarize information.

Language models have also been trained on medical texts, which means they also contain medical knowledge. As a result, they can be used in various ways in the medical sector, including in general practice. Possible applications include writing or analyzing medical letters, answering medical questions, or summarizing medical records. We want to investigate for which tasks you think such a model could be useful, and for which it could not.

From now on, we will refer to these models as "language model" or "AI model". By this, we mean models such as ChatGPT.

**Prompt 2B – Explanation of categories and tasks**

For this study, we have compiled a list of tasks based on the scientific literature and previous conversations with general practitioners. The tasks on this list are divided into five categories: 1) adminstrative tasks 2) pre-consultation and triage 3) clinical decision support 4) management of (chronic) diseases 5) patient interaction and communication. We will go through the list category by category. For each category, we would like to hear your opinion on the tasks it contains.

**Prompt 4A – Closing**

We have now come to the end of the interview; I have asked all of my questions. Thank you very much for your time, your openness, and the valuable insights you have shared. The information from this conversation helps us compile and prioritize a list of tasks for language models in general practice. The results of this study will be published in a scientific article.
